# Impact of obstructive sleep apnoea on cardiometabolic health in an ageing population in rural South Africa: Building the case for the treatment of sleep disorders in deprived settings

**DOI:** 10.1101/2020.11.22.20236000

**Authors:** Johanna Roche, Dale Rae, Kirsten Redman, Kristen L Knutson, Malcolm von Schantz, F Xavier Gómez-Olivé, Karine Scheuermaier

## Abstract

**Objectives:** The association between obstructive sleep apnoea (OSA) and increased cardiometabolic risk (CMR) has been well documented in higher-income countries. However, OSA and its association with CMR have not yet been investigated, based on objective measures, in Southern Africa. We measured polysomnography (PSG)-derived sleep characteristics, OSA prevalence and its association with cardiometabolic diseases in a rural, low-income, aging African-ancestry population in South Africa.

**Methods:** Seventy-five participants were recruited. BMI, hypertension, diabetes, dyslipidaemia, and HIV status were determined. A continuous CMR score was calculated using waist circumference (WC), random glucose, HDL-cholesterol, triglycerides, and mean arterial blood pressure. Sleep architecture, arousal index, and apnoea-hypopnea index (AHI) for detection of OSA (AHI≥15) were assessed by home-based PSG. Associations between CMR score and age, sex, socio-economic status (SES), AHI and TST were investigated by multivariable analysis.

**Results:** In our sample (53 women, 66.1±10.7y, 12 HIV+), 60.7% were overweight/obese, 61.3% hypertensive and 29.3% had undiagnosed OSA. Being older (p=0.02), having a greater BMI (p=0.02) and higher WC (p<0.01) were associated with OSA. AHI severity (ß=0.011p=0.01) and being a woman (ß=0.369, p=0.01) were independently associated with a greater CMR score in SES- and age-adjusted analyses.

**Conclusions:** In this ageing South African community with obesity and hypertension, OSA prevalence is alarming and associated with CMR. We demonstrate feasibility of detecting OSA in a rural setting using PSG. Our results highlight the necessity for actively promoting health education and systematic screening and treatment of OSA in this population, to prevent future cardiovascular morbidity, especially among women.

## Introduction

Obstructive sleep apnoea (OSA) is known as a pro-inflammatory factor (Unnikrishnan, Jun, & Polotsky, 2015) associated with increased risk for cardiometabolic diseases (Kasai, Floras, & Bradley, 2012). It is one of the most prevalent respiratory sleep disorders worldwide with a prevalence ranging from 6-17% among the general adult population, according to data from high-income countries (Senaratna et al., 2017).

In low- to-middle income countries such as those in Sub-Saharan Africa, objective measures of sleep and OSA are scarce, partly due to restricted access to health care. Most of the limited number of studies on sleep performed in the region have assessed sleep quality subjectively using questionnaires such as the Pittsburgh Sleep Quality Index (PSQI) (Gómez-Olivé, Rohr, Roden, Rae, & von Schantz, 2018) and those studies measured the risk of OSA using the Berlin (Adewole et al., 2009) or STOP-BANG (Akanbi et al., 2017; Ozoh et al., 2014) questionnaires. Consequently, knowledge regarding prevalence and consequences of sleep disorders in African populations remains limited. There is strong evidence for increased risk of cardiometabolic disease associated with sleep disorders in African-Americans (AA) compared to other ethnicities in the USA. (Egan, Knutson, Pereira, & von Schantz, 2017; Whitesell, Obi, Tamanna, & Sumner, 2018). However, significant differences in genetic (including admixture), environmental and socio-economic circumstances (Micheletti et al., 2020), precludes extrapolating those findings to the situation of Africans living in Africa. Sub-Saharan Africa, and particularly South Africa, is experiencing a profound health transition, characterized by the emergence of non-communicable diseases (NCDs) and the ageing of the population in both rural and urban areas (Mayosi et al., 2009). For instance, the absolute number of cardiovascular (CV) deaths in Sub-Saharan Africa increased by 81% between 1990 and 2013 (Mensah et al., 2015). A study performed in a rural South African population of African-ancestry adults aged 30 years and above reported an alarming prevalence of obesity, dyslipidaemia and hypertension and concluded that 18.9% of women and 32.1% of men had a 20% or higher chance of having a CV event in the next 10 years, according to their Framingham risk score (Alberts et al., 2005).

In South Africa, the current prevalence of HIV infection (13%) is one of the highest in the world (Statistics Republic of South Africa, 2019). HIV, based on what we know from studies in other populations, constitutes an additional risk factor for sleep disorders (Owens & Hicks, 2018) and cardiometabolic comorbidities (Pullinger et al., 2010). Moreover, rural communities represent a group of interest given the limited access to health services and the high prevalence of NCDs (Gómez-Olivé, Rohr, et al., 2018; Statistics Republic of South Africa, 2019). To date, no community-based study assessing the association between objective sleep and cardiometabolic risk, has been conducted in Sub-Saharan Africa. Such studies are warranted to improve the understanding, detection, and treatment of cardiometabolic disorders in this population.

The aims of this cross-sectional study were: first, to measure PSG-derived sleep characteristics and the prevalence of OSA in an ageing African-ancestry population of low socio-economic status (SES) in a rural community in South Africa; second, to compare the clinical, cardiometabolic, and sleep parameters between participants with and without OSA; and third, to assess sleep parameters associated with CMR in this population in age-, SES- and sex-adjusted analyses.

## Materials and Methods

### 1. Participants

The sample was drawn from the HAALSI cohort created in 2015 (Gómez-Olivé, Montana, et al., 2018), which enrolled 5,059 individuals aged 40 years and older living in the rural Agincourt sub-district in Mpumalanga Province, South Africa. All participants belong to the Shangaan ethnic group. Ninety-two individuals completed home-based PSG. We excluded participants with uninterpretable PSG data (n=11) and uninterpretable respiratory parameters (n=6). The final sample included 75 participants (53 women, 22 men). The flow chart of the study is presented in **Figure 1**.

**Figure 1.**
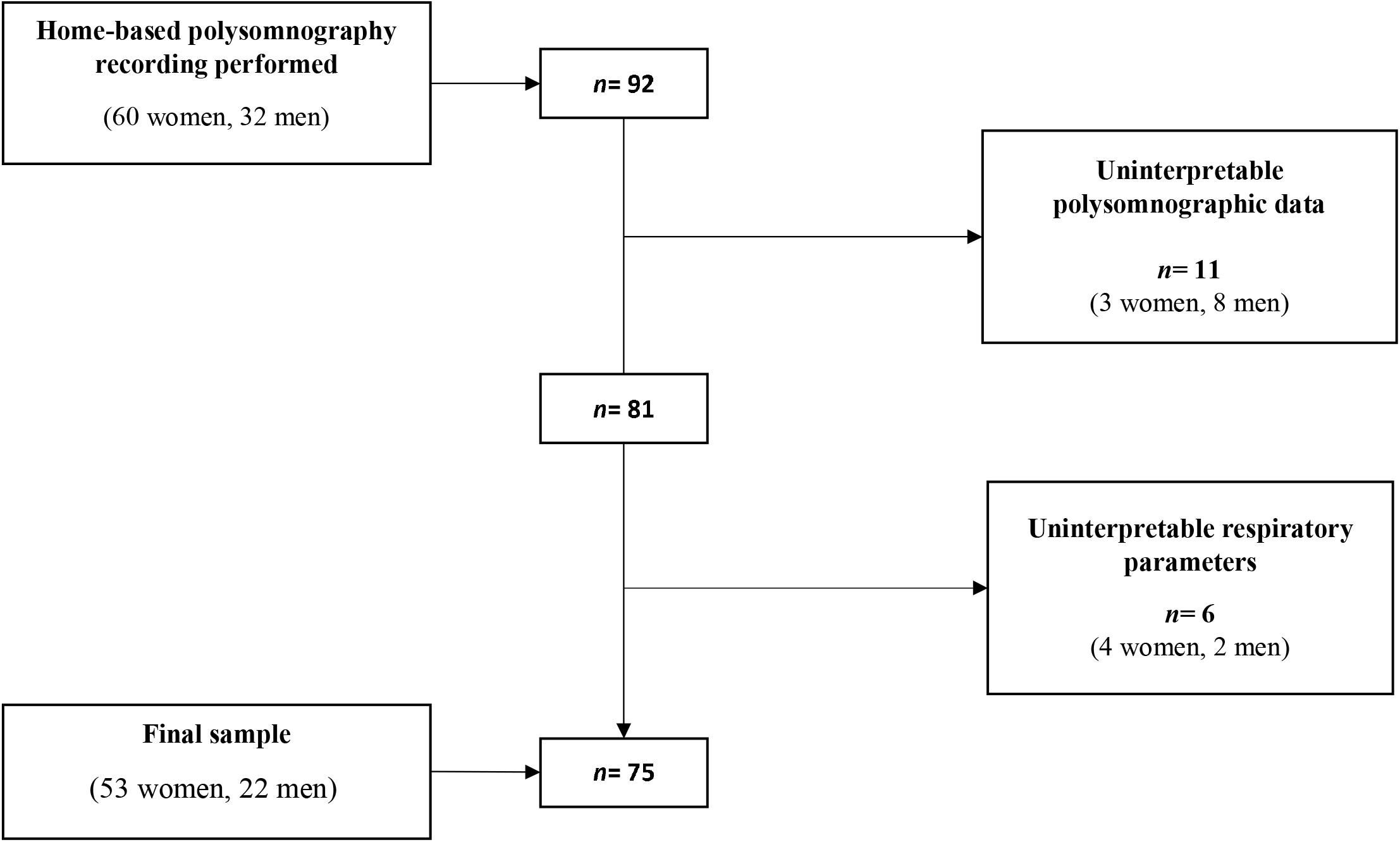
Flowchart of the study population.

### 2. Study design

This is a cross-sectional, observational study in which participants were interviewed in their home by a trained fieldworker. As part of the HAALSI study, demographic information including socio-economic status (SES), relevant personal history of cardiovascular and metabolic disease was collected and anthropometric parameters, blood pressure (BP), and random glucose, lipids, and HIV status were measured. A trained nurse and a locally trained senior fieldworker performed the home-based PSG.

This investigation conformed to the tenets of the Declaration of Helsinki and was approved by the University of the Witwatersrand Human Research Ethics Committee (#M180667) and by the Mpumalanga Province Research Ethics Committee. Written informed consent was obtained from all participants. For illiterate participants, a witness not linked to the study was present during the consenting process.

## 3. Experimental procedures

### 3.1 Demographics

Age and sex were recorded. Household SES level of the participants was determined using a validated SES index, including data on household asset indicators in a typical rural South African setting, from the Agincourt Health and Demographic Surveillance System, over the period 2001-2013 (Kabudula et al., 2017). Full methodology can be found here (Kabudula et al., 2017). This index is presented in raw values and spans from 0 to 5, a lower value indicating lower SES level.

### 3.2 History of cardiometabolic diseases

Participants were asked whether they had had a diagnosis or were/had been under treatment for the following diseases: angina, stroke, heart attack, diabetes, or hypertension. They were classified as having a history of CV event if they reported at least one of the following events: diagnosis of angina, stroke, or heart attack. Classifications of diabetes, dyslipidaemia and hypertension were established according to the blood glucose, lipid and BP measures, and the participants’ declaration of treatment (see sections 3.4 and 3.5).

### 3.3 Anthropometric evaluation

Body weight and height were measured to calculate BMI (kg.m^-2^). According to the World Health Organization recommendations, body weight status was categorized as follows: underweight was defined as BMI< 18.5 kg.m^-2^; normal-weight as BMI ≥ 18.5 but < 25 kg.m^-2^; overweight as BMI ≥ 25 but < 30 kg.m^-2^; and obese as BMI ≥ 30 kg.m^-2^. Waist circumference (WC) was measured in a standing position with a standard non-elastic tape that was applied horizontally midway between the last rib and the superior iliac crest.

### 3.4 Blood pressure measurement

Systolic (SBP) and diastolic (DBP) blood pressures were measured in a seated position after 20min rest using a blood pressure cuff adapted to the arm circumference (Omron M6W® automated cuff, Kyoto, Japan). Measurement was repeated three times at 2-minute intervals and the average of the second and third measurements was used to calculate SBP and DBP. Hypertension stage 2 was defined as a SBP ≥ 140 mmHg, or a DBP ≥ 90 mmHg, or when participants reported current use of medication to treat hypertension (Whelton et al., 2018). Mean arterial pressure (MAP) was calculated using the following formula [(DBP*2) + SBP]/3.

### 3.5 Biochemical measures

Blood was collected using a finger prick for dried blood spots (DBS) for HIV. Point of care measurements were taken for glucose (CareSense, De Puy Synthes, Rainham, MA) and lipids (Cardiochek, PTS Diagnostics, Indianapolis, IN). Participants were classified as diabetic if their non-fasting glucose concentration was ≥11.1 mmol/L or if they reported using medication to control diabetes (Association, 2020). Participants were classified as having dyslipidaemia if they had elevated total cholesterol (≥6.21 mmol/L), or low HDL-C (<1.19 mmol/L), or elevated LDL-C (>4.1 mmol/L), or elevated triglycerides (TG) levels (>2.25 mmol/L), or reported current use of dyslipidaemia treatment at the time of the interview (Gómez-Olivé, Rohr, et al., 2018). HIV status was determined by testing DBS for HIV antibodies with enzyme-linked immunosorbent assays [ELISA; Vironostika Uniform 11 (BioMérieux, Marcy-l’Étoile, France)] and confirmed using the Elecsys® HIV combi PT assay (Roche Diagnostics, Basel, Switzerland) and the ADVIA Centaur® HIV Ag/Ab Combo assay (Siemens Healthcare Diagnostics, Norwood, MA).

### 3.6 Home-based PSG

All participants underwent a single home-based PSG study on a weekday (Track-It Mk2®, Nihon Kohden, Tokyo, Japan). Sleep was assessed using the 10-20 system (JASPER, 1958) and the following electrode derivations were measured and recorded: Fz, Cz, F4-M1, C4-M1, O2-M1, F3-M2, C3-M2 and O1-M2, left and right electrooculogram, and chin electromyogram. Respiratory efforts were measured using thoracic and abdominal inductance plethysmography. Airflow was measured with a nasal pressure cannula. Peripheral oxygen saturation (SpO_2_) was recorded by pulse oximetry (Nonin Medical, Plymouth, MN, USA). The electroencephalogram (EEG) recordings were visually scored in 30-second periods by the first author, using the American Academy of Sleep Medicine’s (AASM) standard rules to obtain the overnight pattern of sleep stages (Berry et al., 2012). We extracted: Sleep latency (time from lights out to sleep defined as the first epoch of any sleep stage, min), total sleep time (TST, h), lights out and lights on (hh:mm, lights out determined as alpha rhythm appearance and lights on as being awake and active), time in bed (lights out to lights on, h), sleep efficiency (TST/time in bed, %), wake after sleep onset (WASO, min), arousal index (events/h of TST), arousal with respiratory events (events), awakening index (events/h of TST), percentage of non-rapid eye movement (NREM) stage 1 sleep in TST (N1, %), percentage of NREM stage 2 sleep in TST (N2, %), percentage of NREM stage 3 sleep in TST (N3, %) and percentage of rapid-eye movement sleep stage in TST (REM, %) and REM latency (min).

Respiratory events (obstructive apnoea (OA), central apnoea (CA), mixed apnoea (MA) and hypopnea) were scored in 3-min periods for airflow according to the adult criteria of the AASM (Berry et al., 2012). OA, CA, MA, hypopnea and oxygen desaturation ≥ 3% index (OAI, CAI, MAI, HI and ODI respectively; events/h of TST) were determined. AHI and ODI were calculated in TST, REM and NREM. Mean SpO_2_, minimum SpO_2_, time spent between 71-80% of SpO_2_, between 81-90% and between 91-100% of SpO_2_ (%) were reported. OSA was defined by the presence of an AHI ≥ 15 events/h of TST.

### 4. Cardiometabolic risk score (CMR score)

A continuous CMR score was calculated in the whole sample of participants (n=92), as described previously (Roche et al., 2020). *Z*-scores of glucose, TG, HDL-C, WC, and MAP average were calculated. Each z-score was obtained by subtracting the sample mean from the individual value divided by the standard deviation (SD) of the sample mean: z-score= (individual value – sample mean)/SD. Then fasting glucose, TG, WC, and MAP z-scores were summed and HDL-C z-score was deducted because of its decreased health risk with higher values. The final value was then divided by 5 to create the continuous CMR score.

### 5. Statistical analyses

Statistical analyses were performed using GraphPad Prism 8.4.3. The Kolmogorov-Smirnov test was used to test the assumption of distribution normality for quantitative continuous parameters with a level of significance set at p<0.05 to reject normality. Data are presented as mean ± standard deviation (SD) for parametric data, median [25-75% Interquartile range (IQR)] for non-parametric data, or *n* (%) for qualitative data. We first compared the OSA and non-OSA groups, then investigated the association between cardiometabolic variables and OSA severity markers in sex, SES- and age-adjusted analyses. Because we found a main effect of sex on several cardiometabolic variables, we also compared men and women for their main demographic, sleep and cardiometabolic characteristics.

Comparisons of clinical, cardiometabolic and sleep variables between participants with and without OSA, and between women and men were performed using unpaired t tests (parametric data) or Mann Whitney tests (non-parametric data). Fisher’s exact tests were used for comparison between categorical variables.

To investigate the association between cardiometabolic variables and OSA severity markers, we first performed linear univariate analyses between the cardiometabolic dependent variables (namely CMR score, glucose, HDL-C, LDL-C, TG, MAP, WC and BMI) and the following independent variables: age, sex, SES, AHI, ODI, arousal index, TST and HIV status. We chose those specific variables (AHI, ODI, arousal index, TST, and HIV status) as they are traditionally associated with a higher cardiometabolic risk (Knutson, 2010; Lavie, 2015; Samaras, Gan, Peake, Carr, & Campbell, 2009; Unnikrishnan et al., 2015). We did not pursue multivariable analysis if we found no significant association between the independent variables and the cardiometabolic dependent variable of interest. We included in our multivariable model analysis independent variables whose univariate’s *p* was <0.2 (to allow for possible confounding effects), and further adjusted for SES, sex, and age.

## Results

Our analysis includes 75 participants (53 women, 22 men) with a mean age of 66.1 ± 10.7 years and a median SES of 2.83 [2.46-3.027]. **Table 1** shows the general and cardiometabolic characteristics of all participants and PSG-derived sleep parameters are shown in **Table 2**. Average TST was 6.57 ± 1.03h, with 14.0% [12-18] of TST classified as N1, 45.2 ± 8.4% as N2, 19.3 ± 8.4% as N3 and 19.6 ± 5.5% as REM. Median AHI was 7.6 [4.3-16.7]. Twenty-two participants had an AHI ≥15, constituting the “OSA group”.

**Table 1.** Demographics and cardiometabolic risk characteristics of all participants and comparison between participants with and without OSA.

|  | **All**<br>**(n= 75)** | **Non-OSA**<br>**(n= 53)** | **OSA**<br>**(n= 22)** | **p** |
| --- | --- | --- | --- | --- |
| **Demographics** |  |  |  |  |
| Age, years | 66.1 ± 10.7 | 64.3 ± 10.9 | 70.4 ± 8.9 | **0.02** |
| Men/women, n (% men) | 22/53<br>(29.33) | 15/38<br>(28.30) | 7/15<br>(31.18) | 0.76 |
| Socioeconomic status score | 2.83 [2.49-3.03] | 2.79 [2.42-3.02] | 2.90 [2.71-3.10] | 0.18 |
| **Cardiometabolic parameters** |  |  |  |  |
| BMI, kg.m<sup>-2</sup> | 28.9 ± 7.4 | 27.6 ± 6.7 | 31.8 ± 8.3 | **0.03** |
| WC, cm | 96.8 ± 18.8 | 93.0 ± 16.2 | 105.4 ± 21.3 | **0.01** |
| Body weight status |  |  |  | 0.20 |
| Underweight, n (%) | 5 (6.8) | 5 (9.6) | 0 (0.0) |  |
| Normal-weight, n (%) | 24 (32.4) | 18 (34.6) | 6 (27.3) |  |
| Overweight, n (%) | 17 (22.9) | 11 (21.2) | 6 (27.3) |  |
| Obese, n (%) | 28 (37.8) | 18 (34.6) | 10 (45.4) |  |
| SBP, mmHg | 139.1 ± 23.6 | 136.1 ± 23.2 | 146.4 ± 23.6 | 0.09 |
| DBP, mmHg | 81.2 ± 11.1 | 80.1 ± 10.7 | 83.7 ± 11.8 | 0.21 |
| Mean Arterial Pressure | 100.5 ± 13.6 | 98.8 ± 13.5 | 104.6 ± 13.4 | 0.09 |
| HTN stage 2, n (%) | 46 (61.3) | 31 (58.5) | 15 (68.1) | 0.60 |
| Controlled, n (%) | 13 (30.2) | 10 (34.5) | 3 (21.4) |  |
| Resistant, n (%) | 8 (18.6) | 4 (13.8) | 4 (28.6) |  |
| Untreated, n (%) | 22 (51.2) | 15 (51.7) | 7 (50) |  |
| History of CV event, n (%) | 8 (10.7) | 8 (15.1) | 0 (0.0) | 0.10 |
| HDL-C, mmol.l<sup>-1</sup> | 1.61 ± 0.47 | 1.72 ± 0.45 | 1.35 ± 0.44 | **<0.01** |
| LDL-C, mmol.l<sup>-1</sup> | 1.97 ± 0.86 | 1.95 ± 0.89 | 2.02 ± 0.83 | 0.76 |
| Triglycerides, mmol.l<sup>-1</sup> | 1.88 ± 1.02 | 1.81 ± 1.00 | 2.04 ± 1.07 | 0.37 |
| Dyslipidaemia n (%) | 30 (42.9) | 17 (34.7) | 13 (61.9) | 0.06 |
| Non-fasting glucose | 5.80 [5.15-7.25] | 5.80 [5.00-7.60] | 6.00 [5.38-6.73] | 0.92 |
| Diabetes, n (%) | 7 (9.6) | 5 (9.8) | 2 (9.1) | 1.00 |
| CMR score | 0.03 ± 0.56 | -0.09 ± 0.57 | 0.29 ± 0.44 | **<0.01** |
| HIV status +, n (%) | 12 (16.7) | 9 (18.0) | 3 (13.6) | 0.74 |

Values are presented as mean ± SD, median [25-75% IQR] or n (%).
BMI= body mass index; WC= waist circumference; SBP= systolic blood pressure, DBP = diastolic blood pressure; HTN= hypertension; CV= cardiovascular; CMR= cardiometabolic risk HDL-C= high-density lipoprotein cholesterol; LDL-C= low-density lipoprotein cholesterol; HIV= human immunodeficiency virus.
Fisher's exact tests, Student T tests or Mann Whitney tests were performed for comparisons between OSA and Non-OSA groups, according to the type and the distribution of the values. For Fisher's exact analysis of body weight status between participants with and without OSA, underweight and normal-weight participants, and overweight and obese participants were grouped.
**Missing data:** body weight status: n=1 (Non-OSA group); information regarding HTN treatment: Non OSA group: n=2, OSA group: n=1; diabetes: n=2 (Non-OSA group); dyslipidaemia: Non-OSA group: n=4, OSA group: n=1; HIV status: n=3 (Non-OSA group).

**Table 2.** Polysomnographic-derived sleep and respiratory parameters of all participants and comparison between participants with and without OSA.

|  | All<br>(*n* = 75) | Non-OSA<br>(*n* = 53) | OSA<br>(*n* = 22) | *p* |
| --- | --- | --- | --- | --- |
| **Sleep parameters** |  |  |  |  |
| Lights out, h:m | 20:48 ± 02:33 | 20:43 ± 03:00 | 20:59 ± 00:39 | 0.69 |
| Lights on, h:m | 05:40 ± 00:56 | 05:40 ± 00:54 | 05:40 ± 01:01 | 0.94 |
| Time in bed, min | 508.8 ± 66.2 | 501.6 ± 69.1 | 526.0 ± 56.9 | 0.15 |
| Sleep latency, min | 10 [5.0-19.8] | 9.0 [5.0-16.0] | 11.0 [5.6-26.4] | 0.39 |
| TST, h | 6.57 ± 1.03 | 6.63 ± 1.08 | 6.42 ± 0.88 | 0.43 |
| Sleep efficiency, % | 81.0 [72.0-85.0] | 82.0 [73.0-87.0] | 75.0 [67.3-82.0] | **0.04** |
| WASO, min | 80.0 [47.3-126.8] | 74.0 [40.0-108.5] | 84.5 [68.3-135.1] | **0.04** |
| N1, % of TST | 14.0 [12.0-18.0] | 13.0 [11.0-17.0] | 15.5 [14.0-20.0] | **<0.01** |
| N2, % of TST | 45.2 ± 8.4 | 46.5 ± 8.5 | 42.0 ± 7.6 | **0.03** |
| N3, % of TST | 19.3 ± 8.4 | 19.2 ± 9.1 | 19.5 ± 6.8 | 0.89 |
| REM, % of TST | 19.6 ± 5.5 | 19.7 ± 6.0 | 19.5 ± 4.1 | 0.89 |
| REM latency, min | 93.8 ± 45.0 | 95.4 ± 44.8 | 89.9 ± 46.3 | 0.63 |
| Total arousals, *n* | 81.0 [50.0-123.5] | 63.0 [48.0-99.0] | 134.0 [92.3-157.0] | **<0.0001** |
| Arousal index, events /h | 14.0 ± 7.3 | 11.4 ± 5.7 | 20.3 ± 7.2 | **<0.0001** |
| Arousal w/ respi event, *n* | 18.0 [8.0-39.5] | 13.0 [6.0-19.0] | 51.5 [39.3-69.5] | **<0.0001** |
| Awakening index, events/h | 4.6 [3.4-5.6] | 4.6 [3.3-5.5] | 4.9 [3.6-5.6] | 0.42 |
| **Respiratory parameters** |  |  |  |  |
| AHI<5, *n* (%) | 27 (36) | 27 (50.9) | - | - |
| 5≤AHI<15, *n* (%) | 26 (34.7) | 26 (49.1) | - | - |
| 15≤AHI<30, *n* (%) | 14 (18.7) | - | 14 (63.6) | - |
| 30≤AHI<45, *n* (%) | 4 (5.3) | - | 4 (18.2) | - |
| AHI≥45, *n* (%) | 4 (5.3) | - | 4 (18.2) | - |
| AHI, events /h | 7.6 [4.3-16.7] | 4.9 [2.9-7.7] | 23.2 [18.7-38.9] | **<0.0001** |
| AHI REM, events /h | 10.5 [4.1-33.2] | 6.4 [1.9-13.6] | 42.6 [30.3-60.8] | **<0.0001** |
| AHI NREM, events /h | 5.9 [3.0-10.9] | 4.0 [1.8-6.3] | 21.2 [14.9-35.5] | **<0.0001** |
| OAI, events /h | 2.9 [0.6-6.6] | 1.2 [0.3-4.0] | 11.0 [5.4-21.8] | **<0.0001** |
| MAI, events /h | 0.0 [0.0-0.0] | 0.0 [0.0-0.0] | 0.0 [0.0-0.2] | **<0.01** |
| CAI, events /h | 0.0 [0.0-0.0] | 0.0 [0.0-0.0] | 0.0 [0.0-0.7] | **<0.01** |
| HI, events /h | 3.8 [2.0-1.8] | 2.7 [1.8-4.7] | 12.8 [11.5-16.4] | **<0.0001** |
| Mean SpO<sub>2</sub>, % | 95.5 ± 2.6 | 96.7 ± 1.5 | 92.9 ± 2.6 | **<0.0001** |
| Min SpO<sub>2</sub>, % | 89.5 [82.8-93.0] | 91.0 [89.0-94.0] | 81.0 [76.0-84.5] | **<0.0001** |
| Total desaturation ≥3%, *n* | 18.2 [3.7-69.0] | 8.7 [1.6-20.1] | 170.2 [71.4-225.3] | **<0.0001** |
| ODI TST 3%, events /h | 2.7 [0.6-11.6] | 1.3 [0.3-3.0] | 25.3 [11.8-31.0] | **<0.0001** |
| ODI REM, events /h | 6.6 [0.0-29.3] | 2.6 [0.0-6.8] | 38.6 [26.6-68.8] | **<0.0001** |
| ODI NREM, events /h | 1.8 [0.4-7.6] | 0.7 [0.2-2.0] | 21.4 [7.9-26.6] | **<0.0001** |
| Time spent between 71-80% SpO<sub>2</sub>, % | 0.0 [0.0-0.0] | 0.0 [0.0-0.0] | 0.0 [0.0-0.8] | **<0.001** |
| Time spent between 81-90% SpO<sub>2</sub>, % | 0.1 [0.0-2.5] | 0.0 [0.0-0.1] | 7.5 [2.4-24.2] | **<0.0001** |
| Time spent between 91-100% SpO<sub>2</sub>, % | 99.5 [93.8-99.9] | 99.9 [98.8-100.0] | 91.9 [58.7-97.5] | **<0.0001** |

Values are presented as mean ± SD or median [25-75% IQR]. TST= total sleep time, WASO= wake after sleep onset, N1= sleep stage 1, N2= sleep stage 2, N3= sleep stage 3, REM= rapid-eye movement sleep, NREM= non rapid-eye movement sleep, AHI= apnoea-hypopnea index, OAI= obstructive apnoea index, CAI= central apnoea index, MAI= mixed apnoea index, HI= hypopnea index, ODI= oxygen desaturation index, Min= minimum, SpO<sub>2</sub>= peripheral oxygen saturation. Unpaired Student T tests, Mann Whitney tests or Fisher's exact test were performed for comparisons between OSA and Non-OSA groups, according to the type and the distribution of the values.

### 1. Comparisons between the OSA and Non-OSA groups

SES status and HIV prevalence were similar between the Non-OSA and OSA groups. Comparison of cardiometabolic characteristics between the two groups is presented in **Table** OSA participants were older (p=0.02), had higher BMI (p=0.02), higher WC (p<0.01) and higher CMR score (p<0.01) than the non-OSA participants.

PSG-derived sleep and respiratory parameters of the two groups are presented in **Table 2**. Participants with OSA had lower sleep efficiency (p=0.04), lower proportion of N2 sleep (p=0.03), higher proportion of N1 sleep (p<0.01), higher arousal index (p<0.001) and more WASO (p=0.04), arousals (p<0.001), and arousals with respiratory events (p<0.001) than the Non-OSA group All respiratory parameters differed between the two groups (all p<0.01).

### 2. Univariate and multivariable linear regressions

In univariate analyses including all participants (*n*=75), CMR score was associated with higher AHI (p=0.004), ODI (p=0.028) and with being a woman (p=0.043, **Table 3)**. We also tested for an interaction between sex and AHI, which was not significant (p=0.30). Therefore, our final multivariable model of CMR score included AHI, SES, age and sex. CMR score was associated with higher AHI (p=0.011), and sex (p=0.011) independently of age (p=0.483) and SES (p=0.548, **Table 3**), whereby higher AHI and being a woman were associated with higher CMR score.

**Table 3.** Predictors of cardiometabolic risk (outcome variable: CMR score) in univariate and multivariable analyses (*n*= 75).

|  | Univariate |  |  | Multivariable |  |  |
| --- | --- | --- | --- | --- | --- | --- |
| | $\beta$ | SE | $p$ | $\beta$ | SE | $p$ |
| Age, years | 0.005 | 0.007 | 0.450 | 0.004 | 0.006 | 0.483 |
| Sex (being a woman) | 0.307 | 0.148 | **0.043** | 0.369 | 0.143 | **0.011** |
| SES | 0.201 | 0.199 | 0.318 | 0.114 | 0.188 | 0.548 |
| AHI, events/h | 0.012 | 0.005 | **0.004** | 0.011 | 0.004 | **0.011** |
| ODI 3%, events/h | 0.013 | 0.006 | **0.028** |  |  |  |
| Arousal index, events/h | 0.004 | 0.009 | 0.684 |  |  |  |
| TST, h | -0.000 | 0.001 | 0.700 |  |  |  |
| HIV status | -0.097 | 0.185 | 0.601 |  |  |  |

AHI= apnoea-hypopnea index, ODI= oxygen desaturation index, TST= total sleep time, HIV= human immunodeficiency virus
Univariate and multivariable analyses were performed using linear regression.

**Supplementary Table 1** shows the results of the univariate and final multivariable analyses performed on the other cardiometabolic variables. In univariate analyses, glucose and MAP were not associated with any of the independent variables considered (all p>0.2). Therefore, no multivariable analysis was pursued. TG was associated with BMI only (p=0.036), but this association was no longer significant after adjustment for age, sex and SES in a multivariable model. LDL-C was associated with age only (p=0.019) in univariate analysis. In the multivariable model, none of the included variables (age, sex, SES, AHI) were associated with LDL-C. HDL-C was inversely associated with AHI (p=0.001), ODI (p<0.01), WC (p<0.0001) and BMI (p<0.001) in unadjusted analyses. In the final multivariable model, HDL-C was negatively associated with AHI (p=0.015) independently of BMI, age, sex, and SES all p>0.05). Higher BMI was associated with higher AHI (p=0.004), ODI (p<0.001), being a woman (p=0.003) and higher SES (p=0.012). In the final multivariable model, BMI was associated with AHI (p=0.004), sex (p<0.0001) and SES (p=0.033) independently of age (p=0.923). Finally, larger WC was associated with higher AHI (p<0.0001), ODI (p<0.001) and SES (p=0.023) in univariate analyses. In the final multivariate model, WC was associated with AHI (p<0.0001) and being a woman (p=0.004) independently of age (p=0.722) and SES (p=0.116).

### 3. Comparisons between the women and men

Since being a woman was associated with several of our cardiometabolic outcome variables, we further compared the demographic, clinical, polysomnography and cardiometabolic characteristics of women and men. This comparison is presented in **Table 4**. Women and men had a similar age and SES. The women had a greater BMI (p<0.001), more overweight/obesity (p=0.02) and a higher CMR score (p=0.04) than the men. Similar proportions of OSA were found among the women (28.3%) and men (31.8%, p=0.76), but men had more N1 sleep (<0.01) and a higher awakening index (p<0.01) compared to the women.

**Table 4.** Cardiometabolic and polysomnographic comparisons between women and men.

|  | Women (*n*= 53) | Men (*n*= 22) | *p* |
| --- | --- | --- | --- |
| **Demographics** |  |  |  |
| Age, years | 65.5 ± 10.8 | 67.5 ± 10.6 | 0.45 |
| Socioeconomic status score | 2.79 [2.42-3.01] | 2.90 [2.72-3.09] | 0.13 |
| **Cardiometabolic parameters** |  |  |  |
| BMI, kg.m<sup>-2</sup> | 30.4 ± 7.6 | 24.9 ± 5.3 | **<0.001** |
| WC, cm | 99.5 ± 19.8 | 90.9 ± 14.8 | 0.08 |
| Body weight status |  |  | **0.02** |
| Underweight, *n* (%) | 2 (3.8) | 3 (14.3) |  |
| Normal-weight, *n* (%) | 14 (26.4) | 10 (47.6) |  |
| Overweight, *n* (%) | 13 (24.5) | 4 (19.05) |  |
| Obese, *n* (%) | 24 (45.3) | 4 (19.05) |  |
| SBP, mmHg | 139.2 ± 23.3 | 138.8 ± 24.9 | 0.94 |
| DBP, mmHg | 82.0 ± 10.8 | 79.2 ± 11.8 | 0.33 |
| Mean Arterial Pressure | 101.1 ± 13.6 | 99.1 ± 13.8 | 0.57 |
| HTN stage 2, *n* (%) | 35 (66.0) | 11 (50.0) | 0.21 |
| Controlled, *n* (%) | 12 (34.3) | 1 (9.1) |  |
| Resistant, *n* (%) | 5 (14.3) | 3 (27.3) |  |
| Untreated, *n* (%) | 18 (51.4) | 7 (63.6) |  |
| History of CV event, *n* (%) | 7 (13.2) | 1 (0.5) | 0.42 |
| HDL-C, mmol.l<sup>-1</sup> | 1.56 ± 0.49 | 1.72 ± 0.43 | 0.21 |
| LDL-C, mmol.l<sup>-1</sup> | 1.86 ± 0.83 | 2.28 ± 0.90 | 0.09 |
| Triglycerides, mmol.l<sup>-1</sup> | 1.91 ± 0.99 | 1.78 ± 1.10 | 0.63 |
| Dyslipidaemia, *n* (%) | 27 (52.9) | 5 (26.3) | 0.06 |
| Non-fasting glucose | 6.0 [5.6-7.4] | 5.6 [4.7-6.8] | 0.06 |
| Diabetes, *n* (%) | 5 (9.4) | 2 (10.0) | 1.00 |
| CMR score | 0.12 ± 0.55 | -0.18 ± 0.54 | **0.04** |
| HIV status +, *n* (%) | 8 (15.7) | 4 (19.0) | 0.73 |
| **Polysomnographic parameters** |  |  |  |
| Lights out, h:m | 20:50 ± 03:00 | 20:41 ± 00:42 | 0.82 |
| Lights on, h:m | 05:40 ± 00:53 | 05:40 ± 01:02 | 0.99 |
| Time in bed, min | 499.2 ± 67.1 | 531.9 ± 59.6 | 0.05 |
| Sleep latency, min | 9.5 [4.5-16.0] | 13.8 [6.1-26.5] | 0.25 |
| TST, h | 6.6 ± 1.0 | 6.6 ± 1.0 | 0.88 |
| Sleep efficiency, % | 82.0 [73.0-85.0] | 75.5 [65.5-84.8] | 0.17 |
| WASO, min | 74.5 [47.0-118.5] | 84.3 [51.0-149.5] | 0.26 |
| N1, % of TST | 14.0 [11.0-16.0] | 18.0 [13.0-21.5] | **<0.01** |
| N2, % of TST | 46.2 ± 7.3 | 42.8 ± 10.4 | 0.11 |
| N3, % of TST | 20.4 ± 8.5 | 16.7 ± 7.8 | 0.08 |
| REM, % of TST | 19.4 ± 5.8 | 20.3 ± 4.7 | 0.49 |
| Arousal index, events /h | 13.1 ± 6.9 | 16.2 ± 8.0 | 0.09 |
| Arousal with respiratory event, *n* | 15.0 [7.0-30.0] | 24.5 [12.0-46.5] | 0.08 |
| Awakening index, events/h | 4.1 [3.0-5.1] | 5.8 [4.5-7.4] | **<0.01** |
| OSA, *n* (%) | 15 (28.3) | 7 (31.8) | 0.76 |
| AHI, events /h | 7.5 [3.9-16.5] | 7.9 [4.6-19.9] | 0.48 |
| Mean SpO<sub>2</sub>, % | 95.6 ± 2.4 | 95.3 ± 3.3 | 0.73 |
| Min SpO<sub>2</sub>, % | 90.0 [82.5-93.0] | 86.0 [83.0-92.0] | 0.87 |
| ODI TST 3%, events /h | 2.7 [0.6-11.1] | 3.4 [1.5-11.6] | 0.69 |

Values are presented as mean ± SD, median [25-75% IQR] or *n* (%).
BMI= body mass index; WC= waist circumference; SBP= systolic blood pressure, DBP = diastolic blood pressure; HTN= hypertension; CV= cardiovascular; CMR= cardiometabolic risk HDL-C= high-density lipoprotein cholesterol; LDL-C= low-density lipoprotein cholesterol; HIV= human immunodeficiency virus, TST= total sleep time, WASO= wake after sleep onset, N1= sleep stage 1, N2= sleep stage 2, N3= sleep stage 3, REM= rapid-eye movement sleep, NREM= non rapid-eye
movement sleep, AHI= apnoea-hypopnea index, OAI= obstructive apnoea index, CAI= central apnoea index, MAI= mixed apnoea index, HI= hypopnea index, ODI= oxygen desaturation index, Min= minimum, SpO<sub>2</sub>= peripheral oxygen saturation.
Fisher's exact tests, Student T tests or Mann Whitney tests were performed for comparisons between OSA and Non-OSA groups, according to the type and the distribution of the values. For Fisher's exact analysis of body weight status between women and men, underweight and normal-weight participants, and overweight and obese participants were grouped.
**Missing data:** body weight status: n=1 (men); diabetes; n=2 (men); dyslipidaemia: women: n=2, men: n=3; HIV status: women: n=2, men: n=1.

## Discussion

In this first objective sleep study in a population of African-ancestry adults living in rural Sub-Saharan Africa, we reported an OSA prevalence of 29.3%. Moreover, OSA severity and being a woman were independently associated with a greater CMR score in SES- and age-adjusted analyses. Overall, our rural population was predominantly female, on average 66 years of age and of low SES. The sex distribution is representative of this region where the low employment opportunities lead to men seeking employment in the bigger cities and therefore the adults remaining are mainly women and /or retired (Gomez-Olive, 2018).

We present some of the first sleep architecture data on sub-Saharan Africans of African ancestry. As a whole group, our participants exhibited similar sleep latency, sleep efficiency, proportions of N3 and REM sleep and arousal index compared to other age-matched populations (Boulos et al., 2019). Time spent in N2 (∼45%) was lower than expected (∼53%), and time in N1 and WASO were greater than normative values for the age (Boulos et al., 2019). This may be partly explained by the elevated prevalence of OSA in our sample. Indeed, our OSA group had lower sleep efficiency, more WASO and N1, less N2 and more arousals than the Non-OSA indicating more fragmented sleep. To date, most of the literature on people of African-ancestry focuses on the comparison between African-Americans, whose genetic background is typically admixed and European-Americans (Ruiter, DeCoster, Jacobs, & Lichstein, 2011; Tomfohr, Pung, Edwards, & Dimsdale, 2012). Those studies found that sleep quality is impaired among African-Americans, with reduced sleep duration, reduced sleep efficiency, and in contrast to our findings, they also found reduced N3 proportion, as well as longer sleep latency, when adjusting for BMI, age, sex and socio-economic status (Ruiter et al., 2011; Tomfohr et al., 2012). Perceived discrimination or habitual living conditions have been incriminated in the altered sleep quality among African-Americans, suggesting that poor sleep might not be only attributable to genetics (Ruiter et al., 2011; Tomfohr et al., 2012). In the present study, our participants underwent PSG in their home. Thus, it seems that sleep quality is not negatively affected by the environment in ageing people living in rural communities. However, given the modest living conditions in rural communities of South Africa [single-room houses, no temperature control indoors, few electric installations (Kahn et al., 2012)], further studies should investigate the possible effects of these parameters on sleep.

Using a standard cut-off of AHI ≥15 events/hour, we found an alarming prevalence of OSA of 29.3%. To date, only two studies reported OSA and risk of OSA prevalence in Sub-Saharan Africa (Ozoh et al., 2014; Poka-Mayap et al., 2020). Ozoh et al. recruited 1100 patients attending a tertiary health facility in Nigeria, with a mean age of 43.9 years. Risk of OSA, as assessed by the STOP-BANG questionnaire, was found in 36.3% of the sample (Ozoh et al., 2014). More recently Poka-Mayap and colleagues assessed OSA prevalence using home sleep testing among patients admitted to a tertiary hospital in Cameroon. Among the 111 participants, with mean age of 58 years, 31.5% had moderate-to-severe OSA as defined using an AHI >15 events/hour (Poka-Mayap et al., 2020). Even though our results are consistent with these two studies, ours is the first to use the gold standard for OSA detection, full polysomnography with recording of respiratory events, in a Sub-Saharan African setting. In contrast with these two studies, we did not recruit in health facilities but used a population-based random sample from an existing older-age cohort (HAALSI). Therefore, our study provides a more accurate estimate of the actual prevalence of OSA in this ageing rural community.

As expected, we found a greater sleep fragmentation and intermittent hypoxia in participants with OSA, expressed by a longer WASO, a greater proportion of N1 and arousals, a lower sleep efficiency and elevated desaturation indices. Both sleep fragmentation and intermittent hypoxia have been involved in the establishment of systemic inflammation, through sympathetic overactivity (Bisogni, Pengo, Maiolino, & Rossi, 2016) and over-production of radical oxygen species pathways (Lavie, 2015), all factors associated with increased cardiometabolic risk.

Chronic inflammation has been reported in treated and untreated HIV infected individuals and has been hypothesized to contribute to the significantly higher risk of cardiovascular disease observed in people living with HIV (PLHW) (Shah Anoop. et al., 2018). A recent study also showed that reporting highly bothersome insomnia was associated with higher risk of cardiovascular events in PLWH over a follow-up period of 10.8 years (Polanka et al., 2019). In this study, we did not measure markers of inflammation and the HIV prevalence (16.7%) was slightly lower than that seen in other studies in the region (23%, Gomez-Olive, 2018) although it was still higher than the national level of 13%. Due to this low representation, we did not have the power to investigate further any interaction between OSA, HIV status and cardiometabolic risk. However, it is likely that the increased activation of inflammatory pathways observed in OSA, may have a more deleterious impact on PLWH, hence contributing more to cardiometabolic risk in the HIV infected population itself.

In the general adult population of higher-income countries, the main identified risk factors for OSA are central obesity, sex, age, heredity and being of African-ancestry (Young, Skatrud, & Peppard, 2004). In our sample, 73% of the participants with OSA were overweight/obese and they exhibited a greater BMI and WC than their counterparts. Additionally, BMI and WC were positively associated with AHI in adjusted multivariate analyses. Ozoh et al. also reported that abdominal obesity, as assessed by WC, was a determinant of nearly double the risk of OSA in a comparable population (Ozoh et al., 2014). Taken together, these results seem to confirm the implication of adiposity, especially central obesity, in the pathogenesis of OSA in this population. Additionally, and in accordance with the literature (Ozoh et al., 2014; Young et al., 2004), older age seems to increase the likelihood of exhibiting OSA. Interestingly, in our sample, men and women were equally affected by OSA, while the literature reports a higher risk of OSA in men (Fietze et al., 2019). However, women exhibited a greater BMI than men, and this factor may explain why OSA is as prevalent in women as in men.

Unexpectedly, we did not find significant cardiometabolic differences between the OSA and Non-OSA groups, apart from the CMR score. For instance, the high prevalence of hypertension and dyslipidaemia were similar in the two groups. Again, these results may be explained, at least in part, by the extremely high prevalence of overweight/obesity found in our entire population. South Africa is still experiencing the emergence of NCDs in both urban and rural areas (Mayosi et al., 2009) and previous surveys conducted in the 2000s already documented the rise of obesity and cardiovascular diseases in rural populations of African-ancestry in South Africa (Alberts et al., 2005; Thorogood et al., 2007). Regarding components of the metabolic syndrome, the CMR score was associated with increased AHI and with the female gender, independently of age and SES. As highlighted before, OSA is a well-established pro-inflammatory factor promoting the emergence of cardiometabolic disorders and, ultimately, endothelial dysfunction (Meier-Ewert et al., 2004; Patel et al., 2009; Unnikrishnan et al., 2015). Regarding sex, and in accordance with previous findings (Gómez-Olivé, Rohr, et al., 2018), we observed a greater overweight/obesity prevalence among women compared to men (69.8% *vs*. 38.1 %, p=0.02).

Accordingly, the present findings call for a better health prevention among rural communities of South Africa where the high prevalence of HIV infection represents an additional cardiometabolic risk factor (Samaras et al., 2009). However, overweight/obesity which promotes the development of OSA, is a modifiable lifestyle factor. A recent multicentric study investigated OSA awareness among primary care physicians in Kenya, Nigeria and in South Africa (Chang et al., 2020). Despite the fact that physicians from South Africa reported a satisfying knowledge regarding OSA, as investigated by the OSA knowledge and Attitudes (OSAKA) questionnaire, overall, respondents showed less confidence in their ability to identify patients at high risk of OSA along with lower confidence in their ability to manage the disorder (Chang et al., 2020). Given the fact that based on our findings, OSA might involve almost one third of the ageing population living in rural South Africa, an effort considering education on sleep and OSA among physicians seems warranted in order to prevent the cardiovascular morbidity/mortality.

This study has some limitations that deserve to be highlighted. Eleven PSG recordings out of the original 92 were uninterpretable. Moreover, an adaptation PSG night was not feasible with our experimental setup. We were also unable to report on any measures of daytime sleepiness or dysfunction, known traits of OSA.

In conclusion, we have provided the first PSG-derived sleep data in a sub-Saharan African cohort of African-ancestry adults and observed an alarming prevalence of undiagnosed and untreated OSA, which was associated with higher CMR. OSA is a modifiable risk factor for cardiometabolic disorders that is currently not screened for or offered treatment for in the public healthcare sector in South Africa, which caters for about 80% of the general population. In a population which is already at high CMR due to traditional risk factors, coupled with a high prevalence of HIV infection, we show that sleep disorders and their treatment may be pivotal aspects of cardiometabolic risk prevention in low-middle-income countries such as South Africa. We highlight the feasibility of performing objective sleep measurements in a deprived rural community using home-based PSG, which then can be used to detect OSA so that treatment can be initiated. Accordingly, it would be worth investing in the introduction of continuous positive airway pressure (CPAP) treatment in participants with OSA in this region and monitoring cardiovascular risk markers during treatment. In the meantime, health and sleep education should be implemented among rural communities, especially among women, and rural health practitioners in order to reduce future cardiovascular morbidity.

## Data Availability

The data will be made available upon reasonable request after publication of the manuscript in a peer-reviewed journal

## Acknowledgements

The authors would like to thank Floidy Wafawanaka, Audrey Khosa, Olivia Khosa and Peter Nkosinathi Tshabangu for their involvement in the data collection. The authors would like to address particular words of thanks to the study participants: *Inkomu Swinene*.

The study was supported by the Academy of Medical Sciences Newton Advance Fellowship to MvS and XGO, and by a postdoctoral fellowship from the University of the Witwatersrand’s University Research Council to JR.

## Author contribution

MvS and XGO designed the study and acquired the funding. DR, KLK and KS helped implement the PSG data collection. JR, DR, KR, MvS, XGO, KS collected the data. JR and KS analysed the data. JR wrote the original draft. DR, KLK, MvS, XGO, and KS reviewed and edited the manuscript.

