## Supplementary Table 1 for "Impact of obstructive sleep apnoea on cardiometabolic health in an ageing population in rural South Africa: Building the case for the treatment of sleep disorders in deprived settings"

**Supplementary Table 1.** Predictors of cardiometabolic markers in univariate and multivariable analyses (*n*= 75).

| **Dependent** | **Univariate** | | | **Multivariable** | | |
| --- | --- | --- | --- | --- | --- | --- |
| **variable** | ***ß*** | **SE** | ***p*** | ***ß*** | **SE** | ***p*** |
| **Mean Arterial Pressure** |  |  |  |  |  |  |
| Age, years | 0,140 | 0,148 | 0.347 |  |  |  |
| Sex (being a woman) | 1,987 | 3,464 | 0.568 |  |  |  |
| SES | 6,433 | 4,472 | 0.155 |  |  |  |
| AHI, events/h | 0,114 | 0,107 | 0.292 |  |  |  |
| ODI 3%, events/h | 0,126 | 0,139 | 0.344 |  |  |  |
| Arousal index, events/h | 0,006 | 0,217 | 0.976 |  |  |  |
| TST, h | 0,007 | 0,026 | 0.769 |  |  |  |
| HIV status | -2,424 | 4,403 | 0.584 |  |  |  |
| BMI, kg.m^-2^ | 0,279 | 0,214 | 0.196 |  |  |  |
| WC, cm | 0,074 | 0,092 | 0.424 |  |  |  |
| **Glucose** |  |  |  |  |  |  |
| Age, years | 0,008 | 0,058 | 0.890 |  |  |  |
| Sex (being a woman) | 1,317 | 1,383 | 0.345 |  |  |  |
| SES | -3,076 | 1,768 | 0.087 |  |  |  |
| AHI, events/h | -0,035 | 0,042 | 0.407 |  |  |  |
| ODI 3%, events/h | -0,356 | 0,367 | 0.341 |  |  |  |
| Arousal index, events/h | 0,020 | 0,084 | 0.816 |  |  |  |
| TST, h | -0,012 | 0,010 | 0.233 |  |  |  |
| HIV status | 0,983 | 1,699 | 0.565 |  |  |  |
| BMI, kg.m^-2^ | -0,036 | 0,084 | 0.668 |  |  |  |
| WC, cm | 0,003 | 0,036 | 0.928 |  |  |  |
| **Triglycerides** |  |  |  |  |  |  |
| Age, years | 0,014 | 0,012 | 0.241 | 0,013 | 0,012 | 0,283 |
| Sex (being a woman) | 0,132 | 0,274 | 0.632 | -0,035 | 0,310 | 0,909 |
| SES | 0,220 | 0,339 | 0.520 | 0,072 | 0,363 | 0,843 |
| AHI, events/h | 0,002 | 0,008 | 0.849 |  |  |  |
| ODI 3%, events/h | 0,005 | 0,010 | 0.660 |  |  |  |
| Arousal index, events/h | 0,009 | 0,016 | 0.566 |  |  |  |
| TST, h | 0,001 | 0,002 | 0.604 |  |  |  |
| HIV status | -0,279 | 0,326 | 0.395 |  |  |  |
| BMI, kg.m^-2^ | 0,034 | 0,016 | **0.036** |  |  | 0.170 |
| WC, cm | 0,010 | 0,007 | 0.149 |  |  |  |
| **LDL-Cholesterol** |  |  |  |  |  |  |
| Age, years | 0,025 | 0,010 | **0.019** | 0,020 | 0,010 | 0,056 |
| Sex (being a woman) | -0,420 | 0,240 | 0.085 | -0,339 | 0,240 | 0,163 |
| SES | 0,492 | 0,319 | 0.128 | 0,338 | 0,319 | 0,294 |
| AHI, events/h | 0,0130 | 0,008 | 0.127 | 0,005 | 0,008 | 0,518 |
| ODI 3%, events/h | 2,904 | 1,876 | 0.129 |  |  |  |
| Arousal index, events/h | 0,021 | 0,014 | 0.141 |  |  |  |
| TST, h | 0,002 | 0,002 | 0.267 |  |  |  |
| HIV status | -0,318 | 0,288 | 0.274 |  |  |  |
| BMI, kg.m^-2^ | 0,017 | 0,015 | 0.250 |  |  |  |
| WC, cm | 0,004 | 0,006 | 0.585 |  |  |  |

**Supplementary Table 1 (continued).** Predictors of cardiometabolic markers in univariate and multivariable analyses (*n*= 75).

| **Dependent** | **Univariate** | | | **Multivariable** | | |
| --- | --- | --- | --- | --- | --- | --- |
| **variable** | ***ß*** | **SE** | ***p*** | ***ß*** | **SE** | ***p*** |
| **HDL-Cholesterol** |  |  |  |  |  |  |
| Age, years | -0,002 | 0,006 | 0.678 | -0,001 | 0,005 | 0,902 |
| Sex (being a woman) | -0,160 | 0,127 | 0.212 | -0,083 | 0,133 | 0,536 |
| SES | -0,135 | 0,159 | 0.399 | 0,042 | 0,155 | 0,786 |
| AHI, events/h | -0,013 | 0,004 | **0.001** | -0,010 | 0,004 | **0.015** |
| ODI 3%, events/h | -10,490 | 3,886 | **0.001** |  |  |  |
| Arousal index, events/h | -0,005 | 0,008 | 0.493 |  |  |  |
| TST, h | 0,001 | 0,001 | 0.261 |  |  |  |
| HIV status | 0,073 | 0,153 | 0.632 |  |  |  |
| BMI, kg.m^-2^ | -0,026 | 0,007 | **<0.001** |  |  |  |
| WC, cm | -0,014 | 0,003 | **<0.001** | -0,018 | 0,009 | 0.053 |
| **BMI** |  |  |  |  |  |  |
| Age, years | 0,038 | 0,081 | 0.644 | -0,007 | 0,070 | 0.923 |
| Sex (being a woman) | 5,563 | 1,814 | **0.003** | 5,697 | 1,603 | **0.001** |
| SES | 5,840 | 2,275 | **0.012** | 4,548 | 2,085 | **0.033** |
| AHI, events/h | 0,182 | 0,055 | **0.001** | 0,156 | 0,053 | **0.004** |
| ODI 3%, events/h | 0,257 | 0,067 | **0.000** |  |  |  |
| Arousal index, events/h | 0,894 | 0,234 | 0.820 |  |  |  |
| TST, h | 0.012 | 0.014 | 0.377 |  |  |  |
| HIV status | -2,381 | 2,476 | 0.340 |  |  |  |
| **WC** |  |  |  |  |  |  |
| Age, years | 0,105 | 0,220 | 0.636 | 0,059 | 0.165 | 0,722 |
| Sex (being a woman) | 8,598 | 4,843 | 0.080 | 11,13 | 3.703 | 0,004 |
| SES | 13,900 | 5,980 | **0.023** | 8,062 | 5.054 | 0,116 |
| AHI, events/h | 0,613 | 0,140 | **<0.001** | 0.601 | 0.1210 | **<0.0001** |
| ODI 3%, events/h | 0,651 | 0,177 | **<0.001** |  |  |  |
| Arousal index, events/h | 0,055 | 0,311 | 0.860 |  |  |  |
| TST, h | 0,024 | 0,037 | 0.523 |  |  |  |
| HIV status | 0,870 | 6,211 | 0.889 |  |  |  |

AHI= apnoea-hypopnea index, ODI= oxygen desaturation index, TST= total sleep time, HIV= human immunodeficiency virus, WC= waist circumference, BMI= body mass index, SES= socioeconomic status. Univariate and multivariable analyses were performed using linear regression.
